# Medical Risk Factors Associated with Listening Difficulties in Children

**DOI:** 10.1101/2024.10.02.24314796

**Authors:** David R. Moore, Adam S. Vesole, Li Lin, Jody Caldwell-Kurtzman, Lisa L. Hunter

## Abstract

**OBJECTIVES:** Listening difficulty refers to difficulty hearing speech despite normal pure-tone audiometry. It is as prevalent as clinical hearing loss among adults, but incidence, causes and treatment remain poorly understood in children. We hypothesized that four medical risk factors would be associated with listening difficulty in children.

**METHODS:** A prospective, case-control study was conducted in a tertiary care children’s hospital. Children (6-13 years old) with clinically normal hearing divided into listening difficulty (n=68) and typically developing (n=84) groups based on a validated caregiver report. All children were native English users without reported conditions restricting participation. Testing included extended high frequency (EHF) audiometry, speech and spatial perception, and cognitive function. Caregiver reports, electronic medical records, and testing ascertained risk of prematurity, head injury, otitis media and EHF hearing loss. Logistic regression, chi-square, correlation, and odds ratios determined associations of listening difficulty with risk factors.

**RESULTS:** Prevalence and risk of prematurity (18%, OR 3.39 [95% CI, 1.1-10.2]), head injury (21%, 3.37 [1.2-9.3]), and high frequency hearing loss (32%, 2.42 [1.1-5.5]) were significantly greater for children with listening difficulty than typically developing children. Ventilation tubes were no more common in the listening difficulty group (25%, 1.14 [0.5-2.4]). EHF hearing loss was associated with prematurity and tubes. Prematurity, tubes, and EHF loss were significantly related to poorer competing speech perception and dichotic listening.

**CONCLUSIONS:** Children with a history of prematurity, head injury or EHF loss were at increased risk of listening difficulties. Early intervention to boost communication skills could potentially improve poorer long-term outcomes.

## INTRODUCTION

Children may present to pediatric or audiology clinics with a primary complaint of listening difficulties (LiD) despite clinically normal audiograms. LiD are usually assessed through validated and reliable self-report questionnaires such as, for adults, the SSQ^1^ or, for children from about 4 years old, caregiver ratings, like the Children’s Communication Checklist (CCC-2^2^) and the ECLiPS^3^. It has been estimated that the number of US adults with LiD, but without hearing loss, is about the same as the number with hearing loss (∼40M in each case^4^). For children, the prevalence of LiD is unknown, but is likely greater than that of hearing loss. Almost all children with LiD have at least one other more recognized neurodevelopmental problem, such as developmental language disorder or attention deficit-hyperactivity disorder (ADHD^5,6,7^), and listening problems are commonly reported in studies of those other disorders^8,9^.

Mechanisms underlying LiD are poorly understood. One possibility is that LiD is a symptom of “sub-clinical” or “hidden” hearing loss. Both terms have been used for several phenomena including elevated tone thresholds outside the clinically normal range of audiometry (0.25 – 8 kHz; ≤ 20 dB HL^10^) and neural temporal summation and/or gain deficits in the cochlea and brainstem^11,12^. Extended high frequency (EHF) hearing plays a role in speech perception in adults^10,13^ and in children^14^. Preliminary evidence of EHF hearing loss (> 8 kHz) has been reported in some children with LiD^15^.

A history of recurrent otitis media (OM), very prevalent in infancy and into the pre-school years^16, 17^, has also been associated with EHF hearing loss^15, 18, 19^, reduced binaural unmasking^20,21^, and increased spatial processing disorder (SPD^22^) in older children. Reported reductions of otoacoustic emissions and wideband middle ear absorbance associated with OM, but increased middle ear reflexes in children with LiD, suggest further possible roles of subclinical hearing loss and altered brainstem function in LiD^23, 24^. However, several studies have found no change in cognition or quality of life following previous pressure equalization (PE) tube insertion^25^, both problems associated with LiD^26^. The specific role of OM in LiD is thus unclear.

Preterm birth and associated neonatal intensive care unit (NICU) stay have been associated with increased incidence of clinical hearing loss^27–29^, and poorer binaural integration and speech-in-noise thresholds later in life^30^. Poorer long-term cognition and language development, and a higher incidence of ADHD^31^ and autism spectrum disorder have all been associated with preterm birth^32^. Given the importance of generalized cognitive performance and association with neurodevelopmental disorders, these outcomes suggest a possible multifaceted impact of preterm birth on LiD, but no such association has been reported in the literature.

Concussions and traumatic brain injury (TBI), collectively “head injury”, are associated with axonal abnormality, demyelination, and elevated extracellular tau levels in the temporal and frontal cortices, as well as subcortically. Both central auditory processing and cognitive components of listening may thus be affected and contribute to LiD^33, 34^. Younger children with histories of head injury have poorer learning and development^35^ and reduced long-term performance in cognition, language and attention^36^. They also have decreased understanding of speech-in-noise and increased within-test fatigue compared to controls^37^. Children with a history of severe TBI have greater caregiver-reported “deafness or problems with hearing” than children with mild TBI^38^ who, in other studies, have not shown significant long-term effects on cognition, language, or attention^39,40^.

No studies have systematically examined the role of these medical risk factors in LiD. Such a study is necessary to provide the earliest possible prevention, detection and intervention for LiD. Previously, we reported that children with LiD had reduced listening, communication, cognitive and auditory skills compared with typically developing children^26, 41^. In this exploratory study of the same cohort, we hypothesized that 1) childhood LiD would associate with medical risk factors of prior OM, EHF hearing loss, head injury and prematurity and, 2) these same medical risk factors would associate with poorer performance on standardized tests of cognition, attention, language, and auditory function markers for LiD.

## METHODS

### Participants

Children (n=152, 6-13 years old at enrollment), were assigned to LiD or typically developing (TD) groups based on their Total ECLiPS score, a reliable and validated caregiver rating scale of childhood listening ability^41–43^. Other inclusion criteria for all participants were English native language, clinically normal hearing bilaterally, and absence of psychiatric, intellectual, or neurologic conditions that would restrict complete testing procedures, determined from caregiver responses to a background questionnaire. TD children additionally had no known history of listening or other recognized neurodevelopmental disorders. Families of participants deemed eligible for the study after preliminary screening completed informed consent, reviewed with the caregiver by a study staff member. Financial compensation was provided to all study families.

### Caregiver Questionnaires

#### Background Questionnaire

Completed by one participant caregiver and included information regarding the participant’s past medical history and demographics. Key variables were birth date, native language, race, ethnicity, parent education, birth history (i.e. weeks premature, length of NICU stay), otologic problems (i.e. OM, PE tube history), developmental or learning difficulties (i.e. attention-deficit disorders, developmental delay, speech-language disorders, autism), TBI or head injury history, history of psychiatric disorders.

#### ECLiPS (Evaluation of Children’s Listening and Processing Skills)

Consists of 38 statements describing stakeholder-elicited descriptions of children’s listening-related behaviors. Caregivers rate on a 5-point scale how strongly they agree or disagree with each statement as it pertains to their child. Statements are divided into 5 subscales: Speech & Auditory Processing, Environmental & Auditory Sensitivity, Language/ Literacy/ Laterality, Memory & Attention, and Pragmatic & Social Skills. Subscales are totaled into 3 composite scores and a Total score, each scaled by age.

#### CCC-2 (Children’s Communication Checklist, 2^nd^ Edition)

Five-point caregiver rating scale that measures a child’s communication skills based on structural and pragmatic aspects of language^2, 44^. It consists of 70 statements with 10 subscales. A General Communication Composite (GCC) score measures clinically significant communication problems while a Social Interaction Difference Index (SIDI) determines the extent of socio-pragmatic difficulties. GCC and SIDI scores are scaled by age.

### Electronic Medical Records (EMR)

A review was made of all 152 participant EMRs (Epic ®) at Cincinnati Children’s Hospital Medical Center. Professional reports of prematurity, TBI or head injury, PE tube insertion, a proxy for recurrent OM^45^, were noted. Caregiver-reported data, where present, were merged with Background Questionnaires and, for prematurity and head injury, analyzed separately from EMRs. Prematurity was defined as birth prior to 37 weeks of gestation. A positive history of PE tubes was either one or more prior EMR reported surgeries or caregiver report.

### Audiometry

Standard clinical audiometry at octave intervals from 0.25 to 8 kHz was performed for each ear. Additional EHF threshold audiometry at 10, 12.5, 14 and 16 kHz was performed for 128 participants.

### Suprathreshold Auditory Tests

#### SCAN-3:C

Age-standardized test battery to assess auditory processing in children aged 5-12 years^46, 47^. Separate tests are: 1) identification of low-pass filtered words, 2) Auditory figure-ground, a speech-in-noise test, 3) Competing words, and 4) Competing Sentences. Competing Words and Sentences involve identification of dichotically presented words or sentences. A Composite score averages across tests.

#### LiSN-S (Listening in Spatialized Noise - Sentences test)

LiSN-S measures a listener’s ability to repeat simple target sentences presented against distracting sentences^48,49^. There are four listening conditions in which the distractors may change talker voice (different or same as target) and/or virtual spatial position (0° or ± 90°). A signal-to-noise ratio at which 50% of sentences are repeated correctly is either the Low Cue speech reception threshold (SRT; same talker, 0° relative to listener) or High Cue SRT (different talker, ± 90° relative to listener). Other LiSN-S scores, obtained by subtracting Low Cue SRT from the different talker SRT or the different position SRT, are the Talker Advantage, and Spatial Advantage, respectively. Total Advantage is the difference between the High Cue and Low Cue SRT. SPD was calculated as both a continuous variable (Pattern score = Spatial Advantage + (Total Advantage - Talker advantage)/2) and a dichotomous variable (presence or absence based on cutoff for SPD = 4.16 + (0.06Age).

### Cognitive Tests

#### NIH Toolbox Cognition Battery

Contains up to eight standardized cognitive tests^50,51^. All participants completed four tests: Picture Vocabulary test, Flanker Inhibitory Control and Attention test, Dimensional Change Card Sort test, and Picture Sequence Memory test, averaged to produce an Early Childhood Composite score. Older participants (≥8 years) completed a Fluid Composite sub-battery (6 memory, attention and executive function tests) and a Crystallized Composite sub-battery (2 reading, vocabulary tests).

### Statistical Analysis

Logistic regression, Chi-square, Spearman correlation, and odds ratios determined associations of LiD with each of the evaluated medical risk factors (prematurity, head injury, EHF hearing loss, PE tubes). Two-sample t-tests and Pearson correlations evaluated relations between risk factors and performance on LiSN-S, NIH Toolbox, CCC, SCAN-3, ECLiPS. Given the exploratory design of the study, multiple testing corrections were only partially applied. Logistic regression analysis determined the association of prior OM with SPD.

## RESULTS

Medical risk factors were identified among both LiD and TD groups, but more so in the LiD than in the TD group (Figure 1).

**Figure 1:**
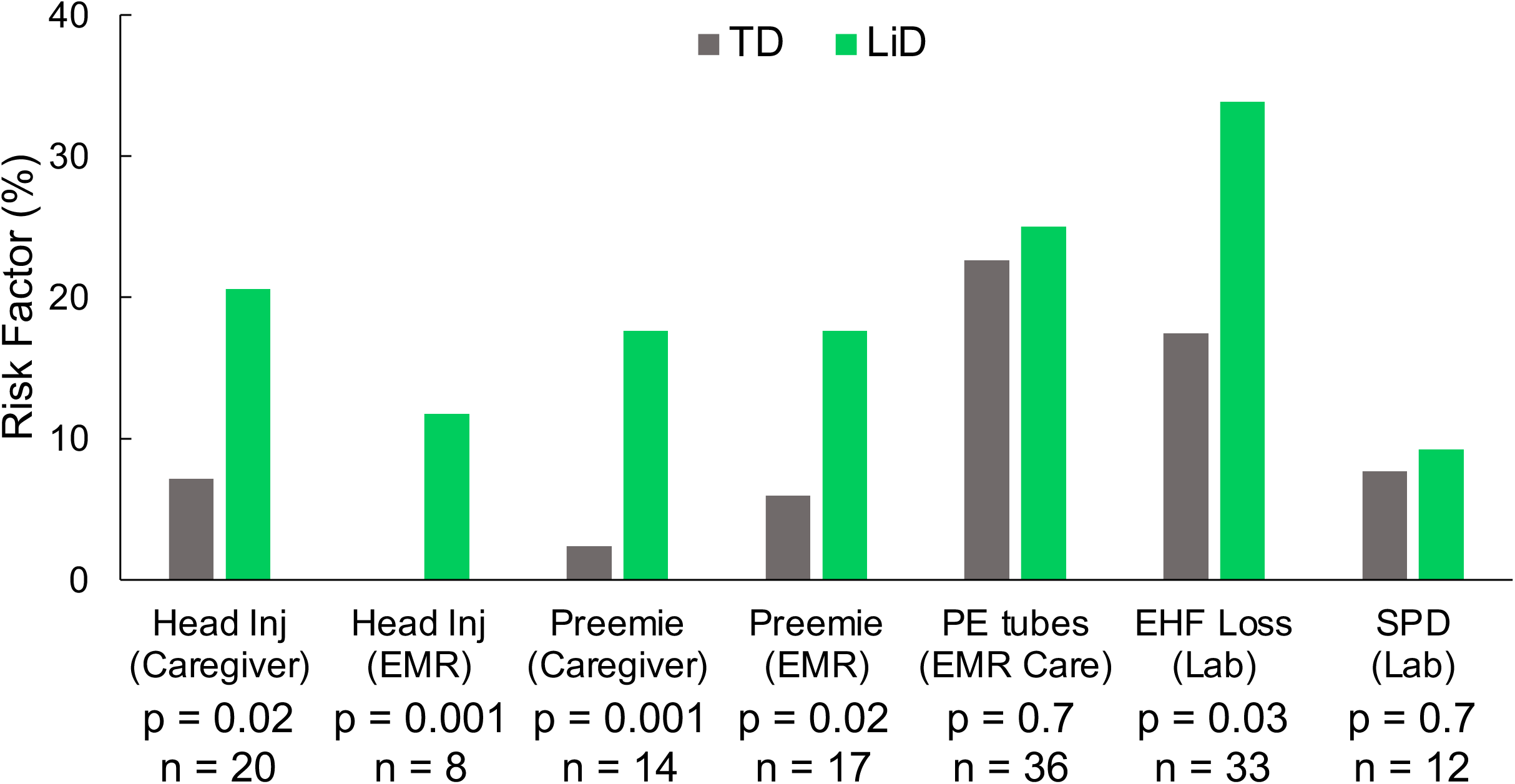
Proportion of children with each medical risk factor in each group (TD – Typically developing, LiD – Listening difficulty), determined by Caregiver report, Medical record (EMR) or Lab testing. Probabilities (p) are based on Chi-squared tests, except Head Injury (EMR) based on Fisher’s Exact test. ‘n’ is the total number of children in both groups for each risk factor.

### Medical risk factors predicted LiD

Statistical analysis of the incidence of each risk factor showed significantly higher rates of prematurity (2-7 weeks early; mean 3.7 weeks), reported head injury, and EHF hearing loss in the LiD group when compared to the TD group (Figure 1, Table 1). Reports of prematurity, head injury, and elevated EHF hearing threshold correlated weakly but significantly with LiD occurrence (Table 2). However, neither PE tube history nor the LiSN-S Pattern measure of SPD were associated with LiD, despite the significant, expected correlation between EHF loss and PE tubes. Rates of caregiver- and EMR-reported prematurity were similar for each group (Figure 1) and were highly and significantly correlated (Table 2), suggesting reliability of caregiver reports. Caregiver- and EMR-reported head injury were more variable, with no cases in the TD group (EMR), and a weaker, but significant correlation. The higher variability was likely due to the subjectivity in classifying a “head injury”. Overall, there was 92% concordance between caregiver-reported and EMR-reported head injury and 99% concordance between caregiver-reported and EMR-reported prematurity, suggesting reliable caregiver recall.

**Table 1:** Logistic Regression, Odds ratio and confirmatory Chi-square Test/Fisher’s Exact Test analysis of the increased risk of childhood LiD, relative to TD children, for each medical risk factor, determined by caregiver report, EMR or laboratory testing. Note that penalized (Firth) logistic regression and Fisher’s Exact Test were applied to the Head Injury EMR due to the small number of cases (n = 8; all in the LiD group).

|  | Prematurity |  | Head Injury |  | PE tubes | EHF Loss |
| --- | --- | --- | --- | --- | --- | --- |
|  | Caregiver | EMR | Caregiver | EMR | EMR and Caregiver | Laboratory |
| Logistic regression | $\chi^2 (1) = 7.7$<br>**p = 0.006** | $\chi^2 (1) = 4.7$<br>**p = 0.03** | $\chi^2 (1) = 5.5$<br>**p = 0.02** | $\chi^2 (1) = 4.2$<br>**p = 0.04** | $\chi^2 (1) = 0.1$<br>p = 0.7 | $\chi^2 (1) = 4.4$<br>**p = 0.04** |
| Odds ratio, 95% CI | 8.8<br>(1.9, 40.8) | 3.4<br>(1.1, 10.2) | 3.4<br>(1.2, 9.3) | 23.7<br>(1.1, 497) | 1.1<br>(0.5, 2.4) | 2.42 (1.06, 5.54) |
| Chi – squared Test | $\chi^2 (1) = 10.5$<br>**p = 0.001** | $\chi^2 (1) = 5.2$<br>**p = 0.02** | $\chi^2 (1) = 6.0$<br>**p = 0.015** | **V = 0.26**<br>**p = 0.001\*** | $\chi^2 (1) = 0.1$<br>p = 0.7 | $\chi^2 (1) = 4.5$<br>**p = 0.03** |

\* Fisher's Exact Test; Cramer's V Bold text: p < 0.05 (uncorrected)

**Table 2:** Spearman’s correlation (ρ) between LiD and medical risk factors. All correlations were calculated from binary (yes/no) contingency matrices. p: probability, n: sample size. Source of data as per Figure 1.

|  | Prematurity<br>Caregiver | Prematurity<br>EMR | Head<br>Injury<br>Caregiver | Head<br>Injury<br>EMR | PE Tubes<br>Caregiver<br>and EMR | EHF loss<br>Lab | SPD<br>Lab |
| --- | --- | --- | --- | --- | --- | --- | --- |
| Listening<br>difficulty | **$\rho = 0.26$**<br>**$p = 0.001$**<br>**$n = 152$** | **0.19**<br>**0.02**<br>**152** | **0.20**<br>**0.02**<br>**152** | **0.26**<br>**0.001**<br>**152** | 0.03<br>0.73<br>152 | **0.19**<br>**0.03**<br>**128** | 0.03<br>0.74<br>143 |
| Prematurity<br>Caregiver<br>Report |  | **0.83**<br>**<.0001**<br>**152** | 0.01<br>0.90<br>152 | 0.03<br>0.74<br>152 | 0.04<br>0.65<br>152 | **0.25**<br>**0.004**<br>**128** | -0.02<br>0.86<br>143 |
| Prematurity<br>EMR |  |  | -0.015<br>0.86<br>152 | 0.01<br>0.90<br>152 | -0.05<br>0.54<br>152 | **0.23**<br>**0.01**<br>**128** | -0.03<br>0.69<br>143 |
| Head<br>Injury<br>Caregiver<br>Report |  |  |  | **0.52**<br>**<.0001**<br>**152** | 0.06<br>0.48<br>152 | -0.05<br>0.61<br>128 | -0.04<br>0.65<br>143 |
| Head Injury<br>EMR |  |  |  |  | -0.06<br>0.45<br>152 | -0.08<br>0.38<br>128 | -0.07<br>0.38<br>143 |
| PE Tubes<br>Caregiver<br>report |  |  |  |  |  | **0.31**<br>**0.0004**<br>**128** | 0.06<br>0.50<br>143 |
| EHF loss<br>Lab |  |  |  |  |  |  | -0.02<br>0.79<br>123 |

Bold text: $p < 0.05$ (uncorrected)

### Risk factors and LiD associated with reduced speech-in-noise and dichotic scores

Two-sample t-tests (Table 3) showed that children with LiD and: 1) caregiver-reported prematurity performed significantly more poorly on multiple outcomes than TD children, 2) prematurity on EMR had significantly poorer scores only for a dichotic, Competing Sentences task, 3) both caregiver- and EMR-reported head injury had significantly poorer scores on two caregiver report outcomes (Communication and Listening), 4) EMR head injury also had significantly poorer scores for dichotic Competing Words, while caregiver-reported head injury had poorer scores on the LiSN-S Talker Advantage task, 5) a history of PE tubes had a significantly poorer score only for the LiSN-S Low Cue task, and 6) EHF hearing loss had significantly poorer scores for Competing Words and the SCAN Composite. Significant findings had effect sizes ranging from d = 0.32-0.61. High concurrence was seen between ECLiPS and CCC-2 caregiver reports. Note that poorer performance on speech-in-noise among children with LiD may be related to OM without OM being an overall risk factor for LiD. Conversely, many individual test score comparisons between the LiD and TD groups did not differ significantly (Table 3), suggesting that test performance was sometimes or often inconsequential for a particular risk factor.

**Table 3:** Comparison of behavioral outcomes between LiD and TD groups for each medical risk factor. Test statistics with significant two-sample t-test values (t: t-score, p: probability, n: sample size, d: effect size). No NIH-Toolbox scores or SPD Pattern scores differed significantly for any risk factor. n.s.: not significant

|  | Prematurity<br>Caregiver | Prematurity<br>EMR | Head<br>Injury<br>Caregiver | Head<br>Injury<br>EMR | PE Tubes<br>Caregiver<br>EMR | EHF<br>Lab |
| --- | --- | --- | --- | --- | --- | --- |
| *ECLiPS* |  |  |  |  |  |  |
| Total | **t = 2.65**<br>**p = 0.01**<br>**n = 149**<br>**d = 0.43** | n.s. | **2.59**<br>**0.01**<br>**149**<br>**0.42** | **2.43**<br>**0.02**<br>**149**<br>**0.40** | n.s. | n.s. |
| *LISN-S* |  |  |  |  |  |  |
| Low-Cue SRT | n.s. | n.s. | n.s. | n.s. | 2.52<br>0.01<br>141<br>0.42 | n.s. |
| Talker Advantage | 2.12<br>0.04<br>141<br>0.36 | n.s. | 2.17<br>0.03<br>141<br>0.37 | n.s. | n.s. | n.s. |
| Spatial Advantage | n.s. | n.s. | n.s. | n.s. | n.s. | n.s. |
| Total Advantage | n.s. | n.s. | n.s. | n.s. | n.s. | n.s. |
| *SCAN* |  |  |  |  |  |  |
| Competing Words | n.s. | n.s. | n.s. | 2.08<br>0.04<br>140<br>0.35 | n.s. | 2.38<br>0.02<br>120<br>0.43 |
| Competing<br>Sentences | **3.28**<br>**0.001**<br>**139**<br>**0.56** | **2.63**<br>**0.01**<br>**139**<br>**0.45** | n.s. | n.s. | n.s. | n.s. |
| Composite | **2.81**<br>**0.01**<br>**139**<br>**0.48** | n.s. | n.s. | n.s. | n.s. | **2.67**<br>**0.01**<br>**119**<br>**0.50** |
| *CCC-2* |  |  |  |  |  |  |
| General<br>Communication<br>Composite | **3.74**<br>**0.0003**<br>**149**<br>**0.61** | n.s. | 1.98<br>0.05<br>149<br>0.32 | **2.95**<br>**0.004**<br>**149**<br>**0.48** | n.s. | n.s. |
| Social Interaction | n.s. | n.s. | n.s. | n.s. | n.s. | n.s. |

□: 0.05, bold: Benjamini-Hochberg multiple testing adjustment

### PE Tube History and Spatial Processing Disorder

The presence or absence of SPD was determined based on the cutoff score, and the Pattern score (Spatial Advantage + (Total Advantage - Talker advantage)/2) was utilized as a continuous variable for SPD. Logistic regression and Fisher exact tests did not identify any statistically significant association between PE tube history and the presence of SPD. Further, a two-sample t-test did not show any association of PE tubes with SPD pattern score.

## DISCUSSION

Discovery of medical risk factors opens the possibility of early detection and intervention for LiD in children. In this study we found one risk factor, prematurity, that is identified at birth, a second, extended high frequency hearing loss, that may be identified soon after birth using physiologic testing or behaviorally from about 18 months, and a third, head injury, that was reported from infancy up to 7 years of age. Each of these factors associated significantly with, and may therefore predict LiD in children. Two other possible risk factors investigated, early OM (PE tubes) and spatial processing disorder (SPD), were both found in young children, but were not associated with LiD. The concurrence between two caregiver reports (ECLiPS and CCC-GCC) for each risk factor adds validity to these results and supports previous suggestions that listening and language are closely related. These validated parent caregiver measures could screen older (≥ 4 years) children for risk factors, potentially leading to formal evaluation.

### Risk factors

This is the first study to analyze multiple medical risk factors and the first to provide evidence for a biological origin of LiD in the absence of clinical hearing loss. Prematurity is associated with multiple medical risks^52^ almost all of which could affect cortical function, hypothesized to be the most critical mechanism of LiD^53^. The late preterm stage of most participants in the study suggests that recognized risks are likely to be low. They could be much higher in very- or extremely-premature infants^54^. Nevertheless, even late prematurity can lead to poor outcomes, including intellectual and mental health disorders^52^.

Long term effects on listening resulting from early environmental influences are suggested by the data on head injuries. These and other preschool age exposures, such as contagious viruses, have also been assumed to contribute to what is generally thought to be a substantial acquired hearing loss^55,56^ and, presumably, LiD following birth. However, data for LiD other than those shown here are not yet available, and the insensitivity of currently used neonatal screening techniques^54,57^ may have led to underestimated incidence of slight-mild sensorineural hearing loss in infancy.

This study is the first to show that EHF hearing loss in older children (≥ 6 y.o.) is associated with caregiver-assessed LiD. EHF hearing loss (> 8 kHz) is likely more prevalent than lower frequency hearing loss from early in childhood^15,18,58,59^, but the evidence is indirect. A high research priority is to determine if EHF hearing loss is present at birth and, if so, include that as a neonatal risk factor for LiD. From the age when children can reliably perform multifrequency audiometry, EHF should be included as part of the test, as has been suggested for older children and adults^13,14^.

Genetic factors may also contribute to LiD in children though there is minimal literature exploring this topic. Twin studies have demonstrated a substantial genetic influence on listening comprehension scores, when compared to environmental factors^60^, and non-speech-based auditory perception^61^. Genetic variants causing clinical hearing loss have been extensively reviewed in the literature. Mutations, such as *USH2A*, selectively affecting the hair cells at the basal cochlea has been associated with LiD and may contribute to EHF hearing loss^62, 63^. However, no significant link between family history of LiD and incidence of auditory processing disorder was found in one study^64^.

### Limitations

Severe prematurity and neurologic issues were excluded, possibly diminishing the power of the study, but this is also a strength, as mild prematurity and head injuries are common and may be major risk factors for listening difficulties. Prematurity and head injury are also known risk factors for speech language and intellectual impairment, which overlap highly with LiD^5–9^. Confirmatory prospective studies incorporating MRI scanning could help to elucidate the links between LiD, prematurity, and neurologic factors. While frequent otitis media and history of PE tube surgery was not a significant risk factor for LiD by parent report, or of SPD, it was associated with the most difficult subtest of the LiSN-S (low cue speech perception).

Prematurity, head injury and EHF hearing loss were found here to have mild to moderate effect sizes on several test outcomes, However, not all these effects were significant when corrected for multiple testing. In addition, the combined effect of these and other risk factors (e.g. head injury AND prematurity) could not be assessed due to lack of statistical power. A confirmatory study with more specific hypotheses and larger samples could now be justified, based on these results.

## CONCLUSION

Children with a history of prematurity, head injury, and extended high frequency hearing loss are at increased risk for listening difficulties that are associated with significant, long-lasting cognitive problems and increased referral to hospital audiology, pediatric, speech/language, psychology, and occupational/physical therapy services. Early intervention to improve communication skills for children with these risk factors could potentially help reduce poorer outcomes that are known to persist at least through adolescence^26^. Medical risk factors should also be considered in older children referred for assessment of listening difficulties. Despite extensive literature linking early and frequent otitis media with listening difficulties, that factor was not found to relate significantly to parent report or test outcomes in this sample. Validated parent report measures could screen children with prematurity, head injury, or reported concern to identify those in need of formal audiologic and speech-language evaluation.

## Data Availability

All data produced in the present study are available upon reasonable request to the authors

## Conflict of Interest Disclosures

No author has any relevant conflict of interest disclosure to declare

## Funding/Support

All phases of this study were supported by NIH grant DC014078 and by the Cincinnati Children’s Research Foundation. Analysis, interpretation and writing were supported by NIH grant DC018734, and by the NIHR Manchester Biomedical Research Centre.

## Abbreviations

ADHD: attention deficit-hyperactivity disorder
CCC-2: Children’s Communication Checklist
EHF: extended high frequency
EMR: electronic medical records
GCC: General Communication Composite
LiD: listening difficulties
LiSN-S: Listening in Spatialized Noise - Sentences test
NICU: neonatal intensive care unit
OM: otitis media
PE: pressure equalization
SIDI: Social Interaction Difference Index
SPD: spatial processing disorder
SRT: speech reception threshold
TD: typically developing
TBI: traumatic brain injury

## Article Summary

Children with caregiver reported listening difficulties had significantly greater rates of preterm birth, head injury, and high frequency hearing loss than typically developing children.

## What’s Known on This Subject

Children born prematurely, or with postnatal otitis media, subclinical hearing loss, or head injury, are at risk for language and other neurodevelopmental disorders.

## What This Study Adds

Children with clinically normal audiograms, but caregiver reported listening difficulties, had significantly greater rates of preterm birth, head injury, and extended high frequency hearing loss, but not otitis media, than those without listening difficulties.

## Contributors Statement

Prof David Moore conceptualized and designed the study, designed and interpreted the analysis, drafted the initial manuscript, and critically reviewed and revised the manuscript.

Dr Adam Vesole drafted the initial manuscript, designed the analysis, and critically reviewed and revised the manuscript.

Li Lin designed, carried out and interpreted the analysis, and critically reviewed and revised the manuscript.

Jody Caldwell Kurtzman collected data, carried out the initial analyses, and critically reviewed and revised the manuscript.

Prof Lisa Hunter conceptualized and designed the study, interpreted the analysis, and critically reviewed and revised the manuscript.

All authors approved the final manuscript as submitted and agree to be accountable for all aspects of the work.

## Notes

### Competing Interest Statement

The authors have declared no competing interest.

### Funding Statement

All phases of this study were supported by NIH grant DC014078 and by the Cincinnati Childrens Research Foundation. Analysis, interpretation and writing were supported by NIH grant DC018734, and by the NIHR Manchester Biomedical Research Centre

### Author Declarations

Cincinnati Childrens Hospital Medical Center Institutional Review Board

